# Experiences and preferences in Zambia and South Africa for delivery of HIV treatment during a client’s first six months: a cross-sectional survey

**DOI:** 10.1101/2024.06.18.24309119

**Authors:** Nyasha Mutanda, Allison Morgan, Aniset Kamanga, Linda Sande, Vinolia Ntjikelane, Mhairi Maskew, Prudence Haimbe, Priscilla Lumano-Mulenga, Sydney Rosen, Nancy Scott

## Abstract

**Background:** Disengagement from antiretroviral therapy (ART) is highest in the early treatment period (≤6 months after initiation/re-initiation), but low intensity models designed to increase retention generally exclude these clients. We describe client preferences for HIV service delivery in the early treatment period.

**Methods:** From 9/2022-6/2023, we surveyed adult clients who were initiating or on ART for ≤6 months at primary health facilities in South Africa and Zambia. We collected data on experiences with and preferences for HIV treatment.

**Results:** We enrolled 1,098 participants in South Africa (72% female, median age 33) and 771 in Zambia (67% female, median age 32), 38% and 34% of whom were initiating/re-initiating ART in each country, respectively. While clients expressed varied preferences, most participants (94% in South Africa, 87% in Zambia) were not offered choices regarding service delivery. 82% of participants in South Africa and 36% in Zambia reported receiving a 1-month supply of medication at their most recent visit; however, South African participants preferred 2- or 3-month dispensing (69%), while Zambian participants preferred 3-or 6-month dispensing (85%). Many South African participants (65%) would prefer to collect medication in community settings, while Zambian participants (70%) preferred clinic-based collection. Half of participants desired more one-on-one counselling and health information. Most participants reported positive experiences with providers, but long waiting queues were reported by South African participants.

**Conclusions:** During the first six months on ART, many clients would prefer less frequent clinic visits, longer dispensing intervals, and frequent, high-quality counselling. Care models for the early treatment period should reflect these preferences.

**Registration:** Clinicaltrials.gov NCT05454839, Clinicaltrials.gov NCT05454852

## Introduction

Among HIV treatment clients in sub-Saharan Africa, the first six months after treatment initiation or re-initiation poses the greatest risk of disengagement from care (stopping treatment). Dubbed the “early treatment period” [1], this interval accounts for roughly three quarters of all first-year attrition from antiretroviral treatment (ART)[2]. In Zambia in 2018-2022, for example, more than a third of clients (34%) experienced interruptions to treatment (ITT) of 28 days or more during their first three months; another 16% interrupted treatment in months 4-6 [3]. By months 7-12, the proportion of clients experiencing ITT had dropped to 12%, after which it plateaued to about 10% per year. During the same years in South Africa, only 56% of clients remained continuously on ART, without any interruptions >28 days, in their first six months after initiation; by 12 months, this proportion dropped to 44% [4].

One challenge to increasing continuity of care during the early treatment period is that newly initiating or re-initiating clients are not eligible for most of the user-friendly differentiated service delivery (DSD) models that have been developed in recent years. DSD models such as six-month dispensing and adherence groups are client-centered approaches that reduce the burden of clinic visits, bring services closer to clients’ homes, and offer some degree of individual choice in determining how services will be accessed [5]. Eligibility criteria for these “low intensity” models of care usually include at least six months of treatment experience and documentation of a suppressed viral load. During the early treatment period, before these criteria can be met, clients are generally required to make frequent clinic visits and receive shorter-duration medication refills. COVID-19 restrictions precipitated some reductions in required numbers of clinic visits and increases in dispensing intervals[6], and new guidelines in South Africa now allow enrollment in DSD models after 4 months on ART, rather than 6 months [7], but for most clients, the early treatment period remains relatively burdensome and inflexible[8].

Adapting DSD strategies to meet clients’ personal preferences during the early treatment period has been proposed as one solution to high early attrition from care [9]. A first step in doing this is to understand those preferences. In this study, we describe preferences for and experiences of HIV care and treatment among ART clients in the early treatment period in South Africa and Zambia.

## Methods

PREFER [10] was a mixed-methods, prospective study of adult ART clients who were either initiating or reinitiating ART or had initiated ART within the previous 6 months at primary healthcare facilities in South Africa and Zambia. The study, which included a baseline survey, focus group discussions, and medical record follow up, addressed new clients’ characteristics, HIV care histories and experiences, resources, needs, concerns, and preferences to help inform the development of appropriate DSD models for the early treatment period. This paper uses the quantitative data to report on prior experience with HIV care and self-reported preferences for how HIV treatment should be delivered during the early treatment period in both countries.

### Study sites and population

PREFER was conducted at the 12 primary healthcare facilities in Zambia and 18 in South Africa that participate in the SENTINEL study of the AMBIT project [10]. They were selected to provide diversity in location (district and province), setting (rural, urban), client volume, DSD model offerings, and nongovernmental support partners. Further descriptions of the study sites have been reported previously [10].

PREFER enrolled new and recent ART initiators who were not yet eligible for low-intensity DSD models. In both countries, eligibility criteria for DSD enrollment at the time of the PREFER survey included i) at least six months on ART; ii) a documented suppressed viral load at the most recent test; and iii) no uncontrolled chronic conditions or co-morbidities, such as noncommunicable diseases, or opportunistic infections that may otherwise compromise clients’ health [11–13]. Inclusion criteria for PREFER study enrollment included living with HIV, being ≥18 years of age, and initiating or re-initiating ART on the day of study enrollment or within the six-month period preceding study enrollment.

### Recruitment and data collection

Upon arrival at study clinics, clients seeking scheduled or unscheduled HIV treatment services were referred to a study research assistant by clinic staff. The research assistant screened potential participants for eligibility and, for those eligible, conducted the informed consent process. Clients who provided written informed consent were enrolled in the study and administered the baseline survey. The questionnaire contained eight thematic sections, including participants’ demographic characteristics and socio-economic status, HIV treatment history, current HIV care and treatment experience, and preferences for features of treatment delivery. Questions on preferences included visit time and frequency, provider interactions, service location, medication dispensing, counselling and health education. On the assumption that an individual’s prior experience with HIV testing and treatment is an important factor in the outcomes of the early treatment period, PREFER also asked several questions about HIV care history. Questions about clients’ perceptions of clinical care were answered using a 5-point Likert scale, describing the proportion of participants who agreed, disagreed, or neither agreed nor disagreed with a series of statements. The study instrument is included as Supplementary File 1.

As PREFER aimed to describe participants’ self-reported experiences and preferences, not compare outcomes or test a hypothesis, the sample size for each study site was chosen to optimize the use of study resources. For each study site, we aimed to enroll 100 participants per site. However, we found that there were fewer eligible clients than originally anticipated. We enrolled just over 60% of our total target sample.

### Quantitative analysis

We first describe participants’ characteristics at enrolment, experience with HIV care, and self-reported preferences on how HIV treatment should be delivered during the first six months of ART treatment using frequencies and simple proportions. Medians with interquartile ranges are reported for continuous variables. For clients’ perceptions of clinical care, which used the 5-point Likert scale described above, we report the proportion of participants who agreed, disagreed, or neither agreed nor disagreed with each of the statements stratified by time on ART and country. To discern differences between newly initiating clients and those who had been on treatment for up to 6 months at study enrolment, we dichotomized participants as either 1) newly initiating or re-initiating ART or 2) as 6 months on ART, based on self-report. Where relevant, we further stratified the first group into ART naïve (newly initiative) and experienced (re-initiating) participants, also based on self-report. All results are presented stratified by country.

### Ethics statement

Country-specific protocols for the PREFER study were approved by Boston University Institutional Review Board under protocol H-42726 (PREFER-South Africa) and H-42903 (PREFER-Zambia). Both protocols were also approved by the University of Witwatersrand Human Research Ethics Committee under protocols M220440 (PREFER-South Africa) and M210342 (PREFER-Zambia). In addition, the protocol for South Africa was approved by the Provincial Health and Research Committees through each study district’s National Health Research Database. The Zambia protocol was also approved by national ethics governing boards, ERES-Converge IRB (2022-June-007) and the Zambia National Health Research Authority (NHRA000007/10/07/2022).

## Results

### Participants’ characteristics

Between 7 September 2022 and 30 June 2023, we screened 1,116 prospective participants in South Africa and 782 in Zambia and enrolled 1,098 in South Africa (72% female) and 771 in Zambia (67% female) in the study. Over a third of participants in South Africa (38%) and Zambia (34%) were initiating or reinitiating ART on the day of study enrolment; the remainder had been on treatment for 0-6 months.

Table 1 reports the demographic characteristics of enrolled participants, stratified by country. In both countries most participants were in their early 30s and were female. Most South African participants were literate (76%) and had completed primary school (63%). In Zambia 43% of participants reported themselves as literate and half had completed primary school. More than half of South African participants lived in urban areas (54%), while the majority of Zambian participants were rural residents (81%). About half of South African participants said they were unemployed (51%), as did about one third of Zambian participants (35%). Most South African participants had electricity (97%), two thirds had access to piped water at the house, and 70% reported household members have never gone without food. In Zambia, 68% had electricity,39% of access to piped water at the house, and more than half (55%) reported that they sometimes or often go without food. In both countries, the majority of participants said that it would be difficult or very difficult for them to come up with the equivalent of $5-6 for a medical emergency (55% South Africa, 82% Zambia). In both countries, about 60% of participants said that they did not know of any other members of their households who were HIV-positive. For those who did report other household members with HIV, though, a majority indicated that at least one of these was on ART. In both countries, roughly two thirds of participants (South Africa 62%, Zambia 66%) had been on treatment for between 1 day and 6 months on the day of study enrollment, while the rest reported they were initiating ART that day.

**Table 1:** Characteristics of PREFER study participants by country.

| Characteristics (n, %) | South Africa | Zambia |
| --- | --- | --- |
| N | 1,098 | 771 |
| Age, median (IQR) | 33 (27, 41) | 32 (27,40) |
| Female | 786 (72) | 514 (67) |
| Marital status |  |  |
| *Live with a primary partner/spouse* | 347 (32) | 362 (47) |
| *Primary partner/spouse but do not live together* | 505 (46) | 123 (16) |
| *No primary partner/spouse* | 246 (22) | 286 (37) |
| Literacy level |  |  |
| *Read well* | 834 (76) | 335 (43) |
| *Read somewhat* | 218 (20) | 235 (30) |
| *Cannot read* | 46 (4) | 201 (26) |
| Highest level of education |  |  |
| *Primary or less* | 409 (37) | 386 (50) |
| *Secondary* | 525 (48) | 329 (43) |
| *Post-secondary* | 164 (15) | 56 (7) |
| Residence location (based on facility setting) |  |  |
| *Rural* | 506 (46) | 623 (81) |
| *Urban* | 592 (54) | 148 (19) |
| Considers house of current residence to be main house |  |  |
| *Yes* | 735 (67) | 738 (96) |
| *Main house is somewhere else in South Africa/Zambia* | 240 (22) | 33 (4) |
| *Main house is in another country* | 123 (11) | - |
| Employment status |  |  |
| *Formal employment* | 240 (22) | 73 (9) |
| *Informal employment* | 216 (20) | 413 (54) |
| *Unemployed* | 562 (51) | 267 (35) |
| *Student/Trainee* | 80 (7) | 18 (2) |
| Access to electricity in house (yes) | 1060 (97) | 524 (68) |
| Access to piped water |  |  |
| *No* | 58 (5) | 205 (27) |
| *Yes, at house* | 736 (67) | 301 (39) |
| *Yes, at community tap/pipe* | 304 (28) | 265 (34) |
| Frequency of HH members going without food |  |  |
| *Never* | 772 (70) | 260 (34) |
| *Seldom* | 72 (7) | 91 (12) |
| *Sometimes* | 220 (20) | 352 (46) |
| *Often* | 34 (3) | 68 (9) |
| Would have difficulty obtaining 100 Rands/100 Kwacha for medical treatment (yes) | 615 (56) | 629 (82) |
| Household members who have HIV (self-report) |  |  |
| *No other HH members with HIV* | 661 (60) | 453 (59) |
| *One other HH member with HIV* | 322 (29) | 252 (33) |
| *Two or more other HH members with HIV* | 115 (10) | 66 (8) |
| If other HH members have HIV, number known to be on ART |  |  |
| *None* | 64 (15) | 49 (15) |
| *One* | 285 (65) | 227 (71) |
| *Two or more* | 88 (20) | 42 (13) |
| Time on ART at study enrollment |  |  |
| *0 days (initiating or re-initiating on day of enrollment)* | 419 (38) | 261 (34) |
| *1-180 days* | 679 (62) | 510 (66) |

HH, Household

Participants’ characteristics are stratified by time on ART (initiated on day of survey or initiated 0-6 months before survey) and sex in Supplementary Table 1. There were few differences in any characteristics by time on ART. Fewer participants initiating or re-initiating ART on the day of the survey reported knowing of any other household members with HIV.

### Experience with HIV care

About half of participants (45% South Africa, 54% Zambia) reported that they had tested for HIV because of ill health (Table 2). One third (34%) of South African participants and 18% of Zambian participants said they had tested HIV-positive before the day on which they started ART, but only 12% in South Africa and 2% in Zambia self-reported previous use of ART (Table 2).

**Table 2.** PREFER study participants’ prior experience with HIV testing and care.

| Experience, n (%) | South Africa | Zambia |
| --- | --- | --- |
| N | 1,098 | 771 |
| Previously tested positive for HIV before starting ART this time | 378 (34) | 135 (18) |
| Previously exposed to ART (prior to current treatment experience) | 134 (12) | 12 (2) |
| Reason for testing at time of first positive test |  |  |
| *Recommended by healthcare provider* | 239 (22) | 141 (18) |
| *Known exposure or risk* | 121 (11) | 103 (13) |
| *Ill health* | 490 (45) | 413 (54) |
| *Pregnancy/antenatal* | 67 (6) | 27 (4) |
| *Self-initiated/voluntary testing* | 155 (14) | 69 (9) |
| *Other* | 26 (3) | 18 (2) |
| Time between testing positive and treatment initiation |  |  |
| *Same day* | 831 (76) | 597 (77) |
| *Within a week* | 186 (17) | 111 (14) |
| *Month plus* | 72 (7) | 60 (8) |
| *Can't remember* | 9 (1) | 3 (0) |
| How many months of HIV medications did you receive today? |  |  |
| *None* | 31 (3) | 34 (4) |
| *< One month* | 2 (0) | 28 (4) |
| *One month* | 896 (82) | 274 (36) |
| *Two months* | 114 (10) | 59 (8) |
| *Three months* | 49 (4) | 334 (43) |
| *Four months* | - | 4 (0) |
| *Six months* | - | 12 (2) |
| *Other* | 6 (1) | 26 (3) |

In both countries, most participants who were initiating or reinitiating treatment on the day of study enrolment received a one-month supply of medications at that visit (82% South Africa, 36% Zambia). Dispensing intervals increased after the initiation visit in Zambia, however: a majority (57%) of those who had started any time prior to the day of study enrolment received a three-month supply. This change did not occur in South Africa, where 75% of those who had started prior to study enrolment reported still received just a one-month supply (Supplementary table 2).

### Preferences for HIV treatment delivery during the first six months of ART treatment

Table 3 presents study participants’ stated preferences for how they would like to receive care if choices were offered, though few participants in either country (6% in South Africa, 13% in Zambia) said that they had been offered any choices of service delivery locations or dispensing durations. There were no significant or programmatic differences in care preferences by sex or location of residence (urban v rural) (Supplementary Tables 2 and 3).

**Table 3.**
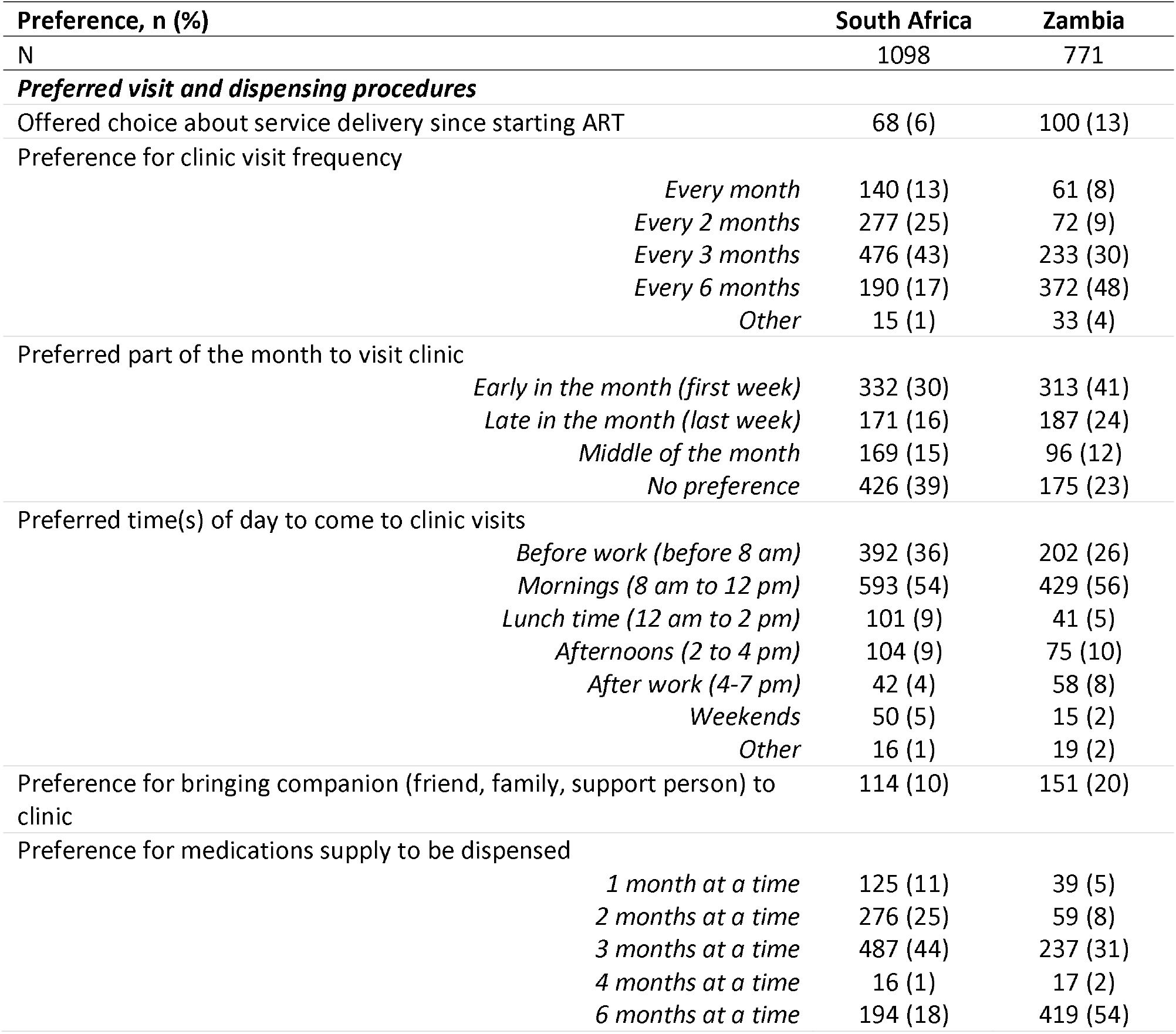
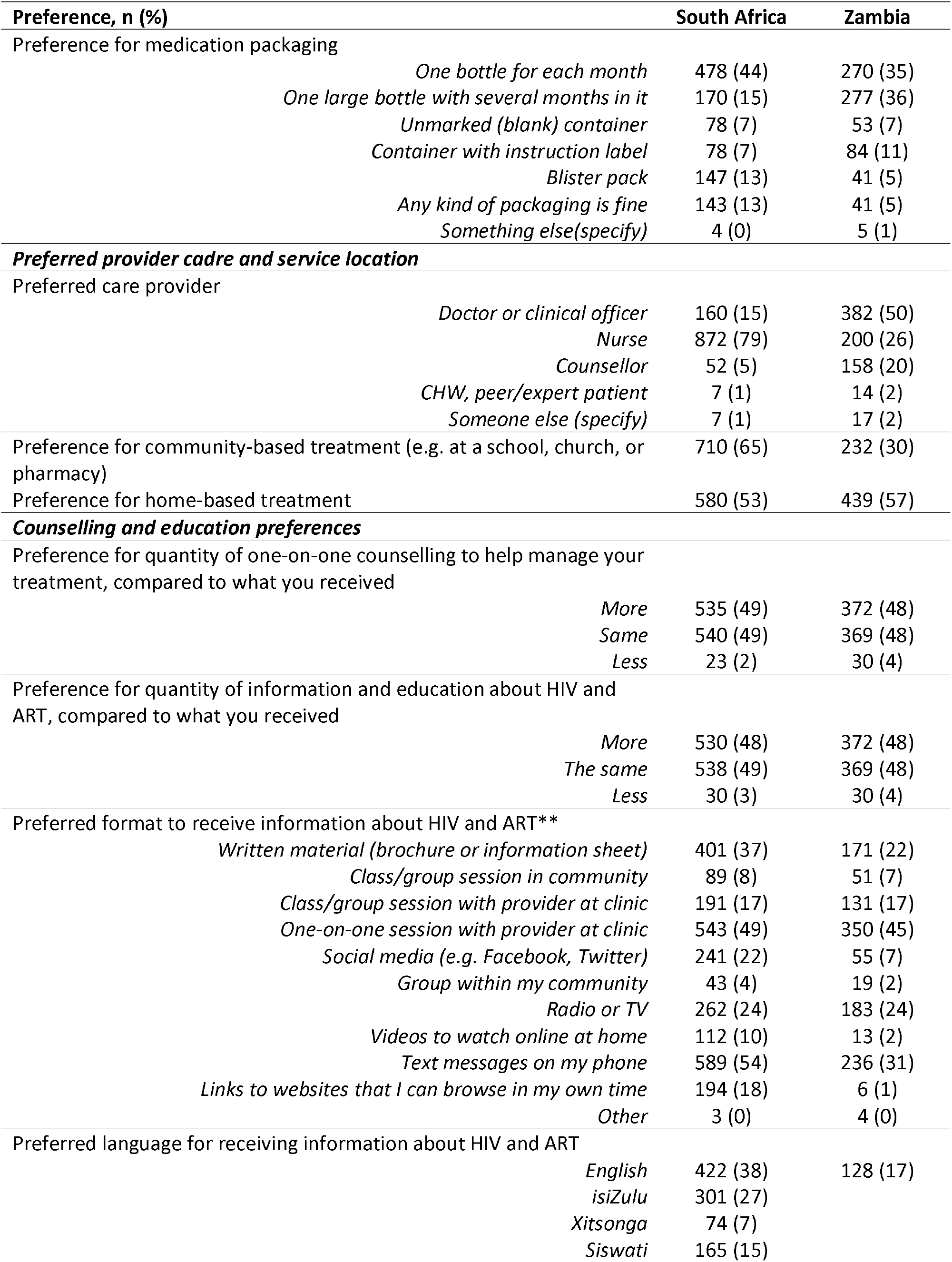
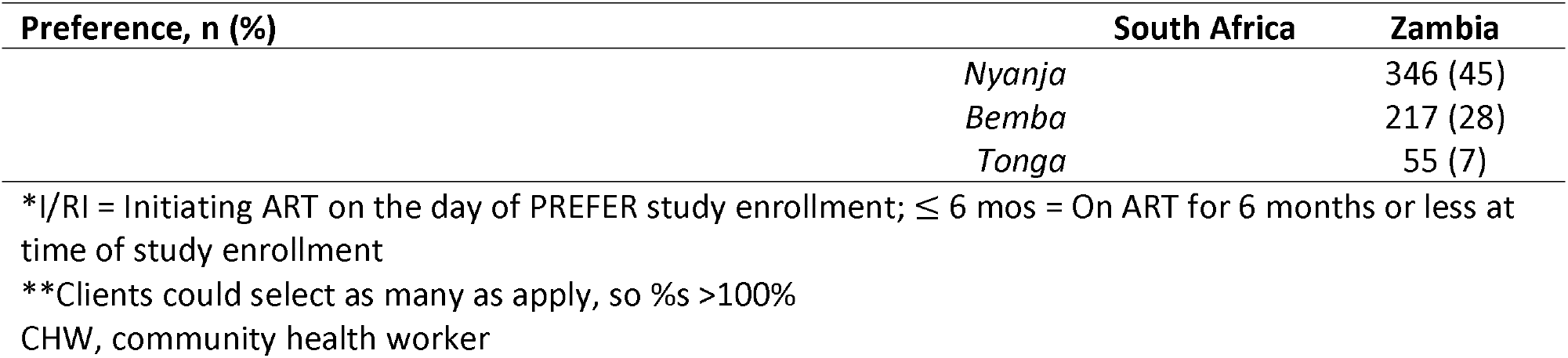
PREFER study participants’ preferences for treatment in the first six months, South Africa and Zambia.

### Preferences for clinic visits and medication dispensing

Clients in South Africa frequently expressed preferences for 2-month (25%) or 3-month (43%) visit frequency; fewer preferred 6-month visits (17%). In contrast, Zambian clients expressed preferences for less frequent visit scheduling, with 48% stating that they would prefer 6-monthly visits. Similarly, clients in South Africa preferred 2 (25%) or 3 months (44%) of medication to be dispensed at a time, while Zambian participants more often preferred 6-month dispensing (54%). Few Zambian participants expressed preferences for shorter 1- or 2-month dispensing intervals. Clients in both countries preferred morning visits (before work or before 12 pm) and most often wanted to visit the clinic alone, instead of with a companion or support person. Participants generally preferred large bottles to store their medication, with either one large bottle for each month, or one bottle to store several months of medication supply.

### Preferred provider cadre and service delivery location

In South Africa, most participants (79%) preferred to receive care from a nurse; in Zambia half (50%) preferred to receive care from a doctor or clinical officer. Very few clients preferred care from a community health worker, peer expert, or expert patient; more Zambian participants (20%) opted for care from a counselor than did South African participants (5%). Most clients in South Africa were receptive to service delivery outside of the health facility, with 65% preferring a community setting, such as a school, church, or pharmacy, and 53% home delivery. Community-based delivery was less preferred in Zambia, where 30% indicated this preference.

### Counselling and health education

About half of participants in both countries expressed a preference for more one-on-one counseling than they said they had received (South Africa: 49%; Zambia: 48%). Most others opted for the same amount of counseling they received; very few wanted less counseling. Trends were similar in both countries when asked about the quantity of information and education they received about HIV and ART: most clients wanted the same or more information. Stratified analysis (Supplementary Tables 2 and 3) showed some variation but few important differences by age group or sex.

Participants varied in their preferences of formats in which to receive information about HIV and ART. In both countries, there were strong preferences for one-on-one sessions with providers in the clinic and informational text messages. In South Africa there was also a preference for written brochures or flyers (37%), while others in Zambia preferred radio/television programming (24%).

### Differences between new initiators and re-initiators

As shown in Table 2, very few Zambian participants (n=12, 2% of sample) voluntarily disclosed previous exposure to ART, but a more substantial proportion in South Africa (n=134, 12%) reported prior treatment experience in the survey responses. In South Africa, other data fields identified an additional 34 South African clients for a total of 168 clients with evidence of prior treatment experience. In Zambia an additional of 30 clients were identified giving a total of 42 clients with previous treatment experience. Table 4 compares study participants initiating or re-initiating treatment on the day of study enrollment who have evidence of prior treatment experience (re-initiators) with those who self-reported as naïve initiators and for whom no other evidence exists. In both countries, absolute numbers of known re-initiators are small, and this should be considered in interpreting the results in Table 4.

**Table 4.** PREFER study participants’ preferences for treatment in the first six months by ART naïve (newly initiating) and experienced (re-initiating) status, South Africa and Zambia.

| Preference, n (%) | South Africa |  |  | Zambia |  |  |
| --- | --- | --- | --- | --- | --- | --- |
|  | Total | Naïve | Reinitiating | Total | Naïve | Reinitiating |
| N (initiators and re-initiators only) | 479 | 311 | 168 | 268 | 226 | 42 |
| ***Preferred visit and dispensing procedures*** |  |  |  |  |  |  |
| Offered choice about service delivery since starting ART | 33 (7) | 14 (5) | 19 (11) | 32 (12) | 21 (9) | 11 (26) |
| Preference for clinic visit frequency |  |  |  |  |  |  |
| *Every month* | 80 (17) | 55 (18) | 25 (15) | 34 (13) | 31 (14) | 3 (7) |
| *Every 2 months* | 120 (25) | 87 (28) | 33 (20) | 32 (12) | 28 (12) | 4 (10) |
| *Every 3 months* | 202 (42) | 127 (41) | 75 (45) | 81 (30) | 64 (28) | 17 (40) |
| *Every 6 months* | 72 (15) | 39 (13) | 33 (20) | 106 (40) | 92 (41) | 14 (33) |
| *Other* | 5 (1) | 3 (1) | 2 (1) | 15 (6) | 11 (5) | 4 (10) |
| Preferred part of the month to visit clinic |  |  |  |  |  |  |
| *Early in the month (first week)* | 137 (29) | 83 (27) | 54 (32) | 109 (41) | 88 (39) | 21 (50) |
| *Late in the month (last week)* | 79 (16) | 50 (16) | 29 (17) | 63 (24) | 56 (25) | 7 (17) |
| *Middle of the month* | 74 (15) | 48 (15) | 26 (15) | 36 (13) | 29 (13) | 7 (17) |
| *No preference* | 189 (39) | 130 (42) | 59 (35) | 60 (22) | 53 (23) | 7 (17) |
| Preferred time(s) of day to come to clinic visits |  |  |  |  |  |  |
| *Before work (before 8 am)* | 184 (38) | 112 (36) | 72 (43) | 70 (26) | 62 (27) | 8 (19) |
| *Mornings (8 am to 12 pm)* | 249 (52) | 170 (55) | 79 (47) | 138 (51) | 113 (50) | 25 (60) |
| *Lunch time (12 am to 2 pm)* | 39 (8) | 24 (8) | 15 (9) | 13 (5) | 10 (4) | 3 (7) |
| *Afternoons (2 to 4 pm)* | 55 (11) | 38 (12) | 17 (10) | 30 (11) | 24 (11) | 6 (14) |
| *After work (4-7 pm)* | 22 (5) | 18 (6) | 4 (2) | 26 (10) | 25 (11) | 1 (2) |
|  | Total | Naïve | Reinitiating | Total | Naïve | Reinitiating |
| *Weekends* | 24 (5) | 20 (6) | 4 (2) | 7 (3) | 7 (3) | 0 (0) |
| *Other* | 9 (2) | 4 (1) | 5 (3) | 6 (2) | 6 (3) | 0 (0) |
| Preference for bringing companion (friend, family, support person) to clinic | 56 (12) | 36 (12) | 20 (12) | 63 (24) | 57 (25) | 6 (14) |
| Preference for medication quantity dispensed |  |  |  |  |  |  |
| *1 month at a time* | 68 (14) | 47 (15) | 21 (12) | 26 (10) | 24 (11) | 2 (5) |
| *2 months at a time* | 121 (25) | 87 (28) | 34 (20) | 26 (10) | 22 (10) | 4 (10) |
| *3 months at a time* | 211 (44) | 134 (43) | 77 (46) | 84 (31) | 67 (30) | 17 (40) |
| *4 months at a time* | 8 (2) | 5 (2) | 3 (2) | 6 (2) | 5 (2) | 1 (2) |
| *6 months at a time* | 71 (15) | 38 (12) | 33 (20) | 126 (47) | 108 (48) | 18 (43) |
| Preference for medication packaging |  |  |  |  |  |  |
| *One bottle for each month* | 226 (47) | 143 (46) | 83 (49) | 87 (32) | 72 (32) | 15 (36) |
| *One larger bottle with several months in it* | 71 (15) | 46 (15) | 25 (15) | 85 (32) | 70 (31) | 15 (36) |
| *An unmarked (blank) container* | 32 (7) | 20 (6) | 12 (7) | 22 (8) | 19 (8) | 3 (7) |
| *A container with instructions on it* | 36 (8) | 27 (9) | 9 (5) | 38 (14) | 32 (14) | 6 (14) |
| *A blister pack* | 57 (12) | 35 (11) | 22 (13) | 23 (9) | 20 (9) | 3 (7) |
| *Any kind of packaging is fine* | 54 (11) | 38 (12) | 16 (10) | 12 (4) | 12 (5) | 0 (0) |
| *Something else* | 3 (1) | 2 (1) | 1 (0) | 1 (0) | 1 (0) | 0 (0) |
| Preferred provider cadre |  |  |  |  |  |  |
| *Doctor or clinical officer* | 83 (17) | 47 (15) | 36 (21) | 125 (47) | 105 (46) | 20 (48) |
| *Nurse* | 369 (77) | 242 (78) | 127 (76) | 64 (24) | 51 (23) | 13 (31) |
| *Counsellor* | 23 (5) | 18 (6) | 5 (3) | 72 (27) | 64 (28) | 8 (19) |
| *Community health worker/peer expert* | 3 (1) | 3 (1) | 0 (0) | 3 (1) | 2 (1) | 1 (2) |
| *Someone else* | 1 (0) | 1 (0) | 0 (0) | 4 (1) | 4 (2) | 0 (0) |
| Preferred location of care |  |  |  |  |  |  |
| *Community-based (e.g. at a school or church or pharmacy)* | 307 (64) | 196 (63) | 111 (66) | 78 (29) | 64 (28) | 14 (33) |
| *Home medication delivery* | 268 (56) | 168 (54) | 100 (60) | 158 (59) | 132 (58) | 26 (62) |
| **Counselling and education preferences** |  |  |  |  |  |  |
| Preference for more/ less one-on-one counselling to help manage your treatment |  |  |  |  |  |  |
| *More* | 250 (52) | 161 (52) | 89 (53) | 135 (50) | 109 (48) | 26 (62) |
| *The same* | 217 (45) | 141 (45) | 76 (45) | 125 (47) | 109 (48) | 16 (38) |
| *Less* | 12 (3) | 9 (3) | 3 (2) | 8 (3) | 8 (4) | 0 (0) |
| Preference for more/less information and education about HIV and ART |  |  |  |  |  |  |
| *More* | 236 (49) | 149 (48) | 87 (52) | 136 (51) | 111 (49) | 25 (60) |
| *The same* | 229 (48) | 151 (49) | 78 (46) | 123 (46) | 106 (47) | 17 (40) |
| *Less* | 14 (3) | 11 (4) | 3 (2) | 9 (3) | 9 (4) | 0 (0) |
| Preferred format to receive information about HIV and ART** |  |  |  |  |  |  |
| *Written material (brochure or information sheet)* | 163 (34) | 112 (36) | 51 (30) | 69 (26) | 55 (24) | 14 (33) |
| *Class/group session in community* | 49 (10) | 32 (10) | 17 (10) | 21 (8) | 19 (8) | 2 (5) |
|  | Total | Naïve | Reinitiating | Total | Naïve | Reinitiating |
| *Class/group session with provider at clinic* | 106 (22) | 61 (20) | 45 (27) | 39 (15) | 32 (14) | 7 (17) |
| *One-on-one session with provider at clinic* | 244 (51) | 160 (51) | 84 (50) | 117 (44) | 105 (46) | 12 (29) |
| *Social media (e.g. Facebook, Twitter)* | 109 (23) | 58 (19) | 51 (30) | 21 (8) | 18 (8) | 3 (7) |
| *Group within my community* | 23 (5) | 15 (5) | 8 (5) | 9 (3) | 8 (4) | 1 (2) |
| *Radio or TV* | 127 (27) | 76 (24) | 51 (30) | 64 (24) | 54 (24) | 10 (24) |
| *Videos to watch online at home* | 45 (9) | 27 (9) | 18 (11) | 2 (1) | 1 (0) | 1 (2) |
| *Text messages on my phone* | 249 (52) | 164 (53) | 85 (51) | 86 (32) | 75 (33) | 11 (26) |
| *Links to websites that I can browse in my own time* | 74 (15) | 51 (16) | 23 (14) | 2 (1) | 2 (1) | 0 (0) |
| *Other* | 3 (1) | 2 (1) | 1 (0) | 3 (1) | 3 (1) | 0 (0) |
| Preferred language for receiving information about HIV and ART, n (%) |  |  |  |  |  |  |
| *English* | 178 (37) | 124 (40) | 54 (32) | 42 (16) | 37 (16) | 5 (12) |
| *isiZulu* | 102 (21) | 65 (21) | 37 (22) | - | - | - |
| *Xitsonga* | 43 (9) | 30 (10) | 13 (8) | - | - | - |
| *Siswati* | 91 (19) | 60 (19) | 31 (18) | - | - | - |
| *Nyanja* | - | - | - | 110 (41) | 90 (40) | 20 (48) |
| *Bemba* | - | - | - | 87 (32) | 74 (33) | 13 (31) |
| *Tonga* | - | - | - | 20 (7) | 16 (7) | 4 (10) |

Differences between naïve and re-initiators were small or non-existent for most preferences. Re-initiators were slightly more likely to report having been given a choice of service delivery characteristics. Preferences for visit and dispensing intervals and timing, medication quantities and packaging, and provider and service location did not differ in any consistent or meaningful way. Re-initiating participants in Zambia were slightly more likely than South African participants to favor more counseling, information, and education than they received, but differences were modest.

### Clients’ views on service delivery

Figure 1 presents survey participants’ reported views on key aspects of HIV treatment delivery at the study sites stratified by country and time on ART (initiating or re-initiating on day of survey enrollment or initiating up to 6 months before survey enrollment). Results were largely but not entirely positive, with regard to views on clinic staff and facilities. Most of the participants in both countries, regardless of their time on ART, strongly agreed that the doctors and nurses had discussed the treatment fully, that they were able to talk with them privately, and that it was easy to tell healthcare providers that they had missed tablets, and most strongly disagreed that providers were too busy to listen to their problems. A minority across both countries and treatment groups, ranging from 10 to 19%, noted difficulty in mentioning missed tablets. More than two thirds of the participants for both treatment groups in South Africa and roughly 40% of those in Zambia strongly or mildly agreed that the queues to see a provider were too long. In Zambia, more than half of the participants disagreed with this statement, generally not perceiving the queues to be too long.

**Figure 1.** Participants’ views of services received in (a) South Africa and (b) Zambia.

## Discussion

In this survey of 1,869 ART clients in South Africa and Zambia during the first six months after initiation or re-initiation of treatment, when they were not eligible for existing DSD models, we found that few choices were offered with regard to the manner of service delivery. This lack of options is consistent with current guidelines, but it may not reflect client preferences and thus may indicate an opportunity for improving early outcomes.

Notably, clients in both countries preferred to receive multi-month medication dispensing and have less frequent visit requirements during the first 6 months on ART. In Zambia, the vast majority of all participants—more than 85%--preferred three or more months of medication dispensed at a time, but 65% actually received only one month at a time. Nearly two thirds of clients in South Africa would prefer 3 or 6 monthly clinic visits with 3-6 months of medications dispensed at a time but, again, guidelines did not allow this for the early treatment period. An unintended consequence of requiring frequent visits and medication refills in the early treatment period is that a client’s early experiences establish their expectations for what treatment will be like going forward. If new clients discover that service delivery is burdensome and inflexible during the early treatment period, they may be deterred from remaining in care until they are eligible for lower-burden DSD models.

Contrary to findings in some other studies among established ART patients in sub-Saharan Africa[14,15], most participants in our survey in both countries reported positive experiences at facilities, particularly with regard to interactions with healthcare providers. Clients generally had positive perspectives of the clinics at which they sought care, expressing trust in the facility, attesting that their providers discussed treatment options with them, and feeling that they had adequate privacy, were comfortable discussing adherence challenges, felt respected, and had their concerns addressed. The reason for this discrepancy between our results and previously published findings is unclear, but it may reflect different provider or client behavior or expectations at initiation and during the early treatment period, compared to later in their treatment journey. It is also possible that the earlier published research revealing the poor regard in which clients held healthcare providers and the importance of better client-provider relationships has in fact improved interactions, and that these improvements are reflected in our more recent data set.

At the same time, many participants commented on long waiting times at facilities and problems retrieving client records, particularly in South Africa. These concerns may reduce motivation to attend future visits [20], especially if visits are required monthly, as is the case in many countries during the early treatment period[8]. In fact, long waiting times were the only facility characteristic that elicited negative reactions from a large minority of study participants.

We were surprised to find that roughly half of participants in both countries expressed a preference for more counseling and information than they received, and virtually none wanted less, despite published doubts about the quality of counseling offered [16]. This was consistent regardless of previous treatment experience and time on ART, suggesting that emotional support and information dissemination are important needs throughout the early treatment period, for all types of clients. Many clients also said that they would like more one-on-one counseling, with nearly half indicating that their preferred method to receive information was through one-on-one sessions with their provider in the health facility. This may present a challenge to improving early treatment outcomes, as anecdotal evidence from treatment providers in both South Africa and Zambia points to reductions in the human resources available to provide counselling and treatment literacy services.

One solution to the discrepancy between the amount and types of counseling that clients reported wanting and what the healthcare system can reasonably offer may be greater use of telehealth options. A substantial proportion of participants in South Africa indicated willingness to receive information through online or remote interaction modalities like social media and links to websites, which could reduce the burden on facility-based services and empower clients to more wholly participate in their care. In Zambia, where internet connectivity is less widespread than South Africa, text messaging support and radio or television broadcasts may be a better approach. While it was once thought that group-based models, such as adherence clubs, would provide the emotional support desired, we speculate that COVID-19 fears and multi-month dispensing, combined with ongoing stigma and fear of disclosure of HIV status, have reduced demand for these models, and most study participants in both countries indicated a preference for individualized counselling over group-based models. Going forward, there may be a renewed role for peer-based support services after ART initiation to replace support that was previously offered by a more robust counseling cadre[17].

In our study, while client preferences were often diverse, stratifying clients by time on ART, gender, and naïve vs. non-naïve status did not result in significant difference in preferences or perceptions of care. Much of the published literature suggests that men’s and women’s care needs and uptake behaviors differ [18,19], but we found that preferences between men and women were not meaningfully different. We also found no important or programmatically meaningful differences in preferences between naïve clients and those re-initiating after a period of disengagement. Individuals within all the key strata—sex, age, prior treatment experience--expressed varied views, with few unanimous or subgroup-specific preferences. While we were able to identify aspects of care that were more or less favored by some subgroups, there were seldom overwhelming majority preferences among our sample.

Based on our findings, we speculate that there other characteristics of subgroups--prior experience with healthcare overall, biases, preconceived notions about HIV, social support, readiness to begin treatment[20], other emotional and personal factors--may affect care preferences more than concrete demographic factors. One implication of this result is that the development of accurate risk stratification approaches to identify individuals at risk of negative outcomes who might require different services, which we believe is a promising direction for further research[21,22], may be even more challenging than expected[24,25]. If self-reported HIV/ART history and basic demographic characteristics do not provide sufficient information to determine optimal service delivery models, the role of self-identification of risk and individual choice of service delivery characteristics may become more important, allowing clients to self-select what is best for them [19,22,23].

Finally, we note that for several of the preferences reported, differences observed between the two countries appear to reflect existing conditions with which clients are familiar. In South Africa, participants preferred two- or three-month intervals between visits and two- or three-month medication dispensing, with care provided by a nurse; this was the standard of care at the time of the survey. In Zambia, where national guidelines allowed 6-month dispensing once a client is eligible for DSD and most clinical consultations are provided by clinical officers, participants said they preferred 6-month dispensing and care from clinical officers. It is thus unclear what clients’ true preferences would be in the absence of clinical norms and experience [15,16].

There were several limitations to this study. The sample size was modest and included only a handful of facilities in each country, potentially limiting generalizability. As with any survey, results are based on self-report, which may or may not reflect actual experience or views. This is particularly important when comparing ART naïve to re-initiating clients, as there is evidence that non-disclosure of prior treatment experience is common [23]. Our ability to distinguish naive and non-naive initiators was poor, and comparisons of these populations must be interpreted with caution. In addition, this study was conducted prior to the adoption of new HIV care guidelines in South Africa. The recently adopted guidelines for South Africa allow eligibility for DSD models as early as four months after treatment initiation for those with a suppressed viral load, rather than the previous six months [7]. Finally, we assume that our results reflect some degree of social desirability bias, leading to more positive responses to some questions than might be truly representative of participants’ views.

## Conclusion

In many countries, the early treatment period remains a challenge for delivering HIV treatment in a way that supports retention in care and other positive outcomes. While we did not identify a “smoking gun” to explain the high rates of interruption and disengagement observed during this period, we did find a wide range of preferences that can be used to improve specific aspects of service delivery for specific subgroups of the population. We also found that the preferences of clients in the early treatment period align with existing DSD models designed for established clients, such as multi-month dispensing in Zambia and community-based delivery (external pickup points) in South Africa. Existing models may thus offer a strong base on which to build more effective models for the early treatment period.

## Data Availability

All data used in the present study are available upon reasonable request to the authors and will be posted in a public repository following the closing of the ethics protocol.

## List of abbreviations

ART: Antiretroviral therapy
DSD: Differentiated service delivery
HH: Household
ITT: Interruptions to treatment

## Supplementary files

Table S1. Participant characteristics by country, sex, and time on treatment at enrolment

Table S2. Participants’ preferences for treatment in the first six months by sex, age, and time on ART at enrolment, South Africa

Table S3. Participants’ preferences for treatment in the first six months by sex, age, and time on ART at enrolment, Zambia

File S1. Survey instrument

## Acknowledgements

Funding for the study was provided the Bill & Melinda Gates Foundation through award INV-031690. The funder had no role in study design, data collection, analysis, or preparation of this manuscript.

## Supplementary Tables

**Supplementary Table 1.** Participant characteristics by country, sex, and time on treatment at enrolment.

| Characteristic (n, %) | South Africa |  |  |  |  | Zambia |  |  |  |  |
| --- | --- | --- | --- | --- | --- | --- | --- | --- | --- | --- |
|  | Total | Sex |  | Time on ART at enrolment |  | Total | Sex |  | Time on ART at enrolment |  |
|  |  | Male | Female | I/RI* | ≤6 mos |  | Male | Female | I/RI* | ≤6 mos |
| N | 1,098 | 312 (28) | 786 (72) | 419 (38) | 679 (62) | 771 | 257 (33) | 514 (67) | 261 (34) | 510 (66) |
| Age, median (IQR) | 33 (27, 41) | 38 (33, 44) | 31 (25, 39) | 34 (27,40) | 33 (27,41) | 32 (27,40) | 36 (30, 43) | 31 (25, 39) | 32 (27,40) | 33 (27,40) |
| Female | 786 (72) |  |  | 289 (69) | 497 (73) | 514 (67) |  |  | 151 (58) | 363 (71) |
| Marital status |  |  |  |  |  |  |  |  |  |  |
| *Live with a primary partner/spouse* | 347 (32) | 131 (42) | 216 (27) | 137 (33) | 210 (31) | 362 (47) | 141 (55) | 221 (43) | 126 (48) | 236 (46) |
| *Primary partner/spouse but do not live together* | 505 (46) | 119 (38) | 386 (49) | 180 (43) | 325 (48) | 123 (16) | 40 (16) | 83 (16) | 40 (15) | 83 (16) |
| *No primary partner/spouse* | 246 (22) | 62 (20) | 184 (23) | 102 (24) | 144 (21) | 286 (37) | 76 (30) | 210 (41) | 95 (36) | 191 (37) |
| Literacy level |  |  |  |  |  |  |  |  |  |  |
| *Read well* | 834 (76) | 223 (71) | 611 (78) | 314 (75) | 520 (77) | 335 (43) | 136 (53) | 199 (39) | 125 (48) | 210 (41) |
| *Read somewhat* | 218 (20) | 72 (23) | 146 (19) | 89 (21) | 129 (19) | 235 (30) | 71 (28) | 164 (32) | 70 (27) | 165 (32) |
| *Cannot read* | 46 (4) | 17 (5) | 29 (4) | 16 (4) | 30 (4) | 201 (26) | 50 (19) | 151 (29) | 66 (25) | 135 (26) |
| Highest level of education |  |  |  |  |  |  |  |  |  |  |
| *Primary or less* | 409 (37) | 131 (42) | 278 (35) | 147 (35) | 262 (39) | 386 (50) | 113 (44) | 273 (53) | 125 (48) | 261 (51) |
| *Secondary* | 525 (48) | 147 (47) | 378 (48) | 213 (51) | 312 (46) | 329 (43) | 119 (46) | 210 (41) | 115 (44) | 214 (42) |
| *Post-secondary* | 164 (15) | 34 (11) | 130 (17) | 59 (14) | 105 (15) | 56 (7) | 25 (10) | 31 (6) | 21 (8) | 35 (7) |
| Considers house of current residence to be main house |  |  |  |  |  |  |  |  |  |  |
| Yes | 735 (67) | 217 (70) | 518 (66) | 279 (67) | 456 (67) | 738 (96) | 247 (96) | 491 (96) | 246 (94) | 492 (96) |
|  | Total | Sex |  | Time on ART at enrolment |  | Total | Sex |  | Time on ART at enrolment |  |
|  |  | Male | Female | I/RI* | ≤6 mos |  | Male | Female | I/RI* | ≤6 mos |
| *No, main house is somewhere else in South Africa/Zambia* | 240 (22) | 58 (19) | 182 (23) | 92 (23) | 148 (22) | 33 (4) | 10 (4) | 23 (4) | 15 (6) | 18 (4) |
| *No, main house is in another country* | 123 (11) | 37 (12) | 86 (11) | 48 (11) | 75 (11) | - | - | - | - | - |
| Employment status |  |  |  |  |  |  |  |  |  |  |
| *Formal employment* | 240 (22) | 108 (35) | 132 (17) | 91 (22) | 149 (22) | 73 (9) | 45 (18) | 28 (5) | 35 (13) | 38 (7) |
| *Informal employment* | 216 (20) | 84 (27) | 132 (17) | 93 (22) | 123 (18) | 413 (54) | 165 (64) | 248 (48) | 135 (52) | 278 (55) |
| *Unemployed* | 562 (51) | 112 (36) | 450 (57) | 201 (48) | 361 (53) | 267 (35) | 43 (17) | 224 (44) | 85 (33) | 182 (36) |
| *Student/Trainee* | 80 (7) | 8 (3) | 72 (9) | 34 (8) | 46 (7) | 18 (2) | 4 (2) | 14 (3) | 6 (2) | 12 (2) |
| Access to electricity in house (yes) | 1060 (97) | 294 (94) | 766 (97) | 405 (97) | 655 (96) | 524 (68) | 157 (61) | 367 (71) | 179 (69) | 345 (68) |
| Access to piped water |  |  |  |  |  |  |  |  |  |  |
| *No* | 58 (5) | 18 (6) | 40 (5) | 20 (5) | 38 (6) | 205 (27) | 68 (26) | 137 (27) | 56 (21) | 149 (29) |
| *Yes, at house* | 736 (67) | 189 (61) | 547 (70) | 283 (68) | 453 (67) | 301 (39) | 94 (37) | 207 (40) | 109 (42) | 192 (38) |
| *Yes, at community tap/pipe* | 304 (28) | 105 (34) | 199 (25) | 116 (28) | 188 (28) | 265 (34) | 95 (37) | 170 (33) | 96 (37) | 169 (33) |
| Frequency of HH members going without food |  |  |  |  |  |  |  |  |  |  |
| *Never* | 772 (70) | 213 (68) | 559 (71) | 295 (70) | 477 (70) | 260 (34) | 102 (40) | 158 (31) | 96 (37) | 164 (32) |
| *Seldom* | 72 (7) | 19 (6) | 53 (7) | 23 (5) | 49 (7) | 91 (12) | 26 (10) | 65 (13) | 27 (10) | 64 (13) |
| *Sometimes* | 220 (20) | 73 (23) | 147 (19) | 91 (22) | 129 (19) | 352 (46) | 111 (43) | 241 (47) | 119 (46) | 233 (46) |
| *Often* | 34 (3) | 7 (2) | 27 (3) | 10 (2) | 24 (4) | 68 (9) | 18 (7) | 50 (10) | 19 (7) | 49 (10) |
| Would have difficulty obtaining 100 Rands/100 Kwacha for medical treatment (yes) | 615 (56) | 170 (54) | 445 (56) | 236 (56) | 379 (56) | 629 (82) | 197 (77) | 432 (84) | 214 (82) | 415 (81) |
| Other HH members who have HIV (self-report) |  |  |  |  |  |  |  |  |  |  |
| *No other HH members with HIV* | 661 (60) | 175 (56) | 486 (62) | 271 (65) | 390 (57) | 453 (59) | 152 (59) | 301 (59) | 180 (69) | 273 (54) |
|  | Total | Sex |  | Time on ART at enrolment |  | Total | Sex |  | Time on ART at enrolment |  |
|  |  | Male | Female | I/RI* | ≤6 mos |  | Male | Female | I/RI* | ≤6 mos |
| *One other HH member with HIV* | 322 (29) | 111 (36) | 211 (27) | 118 (28) | 204 (30) | 252 (33) | 86 (33) | 166 (32) | 69 (26) | 183 (36) |
| *Two or more other HH members with HIV* | 115 (10) | 21 (8) | 89 (11) | 30 (7) | 85 (12) | 66 (8) | 60 (23) | 82 (16) | 12 (5) | 54 (10) |
| If other HH members have HIV,<br>number known to be on ART |  |  |  |  |  |  |  |  |  |  |
| *None* | 64 (15) | 15 (11) | 49 (16) | 20 (14) | 44 (15) | 49 (15) | 15 (14) | 34 (16) | 12 (15) | 37 (16) |
| *One* | 285 (65) | 101 (74) | 184 (61) | 105 (71) | 180 (62) | 227 (71) | 78 (74) | 149 (70) | 62 (77) | 165 (70) |
| *Two or more* | 88 (20) | 21 (15) | 67 (22) | 23 (15) | 65 (23) | 42 (13) | 12 (11) | 30 (14) | 7 (8) | 35 (14) |

**Supplementary Table 2.** Participants’ preferences for treatment in the first six months by sex, age, and time on ART at enrolment, South Africa.

| Preference (n, %) | Total | Sex |  | Age groups (years) |  |  | Place of Residence |  | Time on ART at study enrolment |  |
| --- | --- | --- | --- | --- | --- | --- | --- | --- | --- | --- |
|  |  | Male | Female | 18-24 | 25-49 | 50+ | Rural | Urban | *I/RI | ≤6 mos |
| N | 1098 | 312 | 786 | 188 | 801 | 109 | 506 | 592 | 419 | 679 |
| ***Preferred visit and dispensing procedures*** |  |  |  |  |  |  |  |  |  |  |
| Offered choice about service delivery since starting ART (yes) | 68 (6) | 21 (7) | 47 (6) | 13 (7) | 48 (6) | 7 (6) | 16 (3) | 52 (9) | 31 (7) | 37 (5) |
| Preference for clinic visit frequency |  |  |  |  |  |  |  |  |  |  |
| *Every month* | 140 (13) | 49 (16) | 91 (12) | 25 (13) | 103 (13) | 12 (11) | 61 (12) | 79 (13) | 73 (17) | 67 (10) |
| *Every 2 months* | 277 (25) | 81 (26) | 196 (25) | 49 (26) | 195 (24) | 33 (30) | 129 (25) | 148 (25) | 107 (26) | 170 (25) |
| *Every 3 months* | 476 (43) | 127 (41) | 349 (44) | 74 (39) | 354 (44) | 48 (44) | 228 (45) | 248 (42) | 174 (42) | 302 (44) |
| *Every 6 months* | 190 (17) | 51 (16) | 139 (18) | 39 (21) | 139 (17) | 12 (11) | 78 (15) | 112 (19) | 62 (15) | 128 (19) |
| *Other* | 15 (1) | 4 (1) | 11 (1) | 1 (1) | 10 (1) | 4 (4) | 10 (2) | 5 (1) | 3 (1) | 12 (2) |
| Preferred part of the month to visit clinic |  |  |  |  |  |  |  |  |  |  |
| *Early in the month (first week)* | 332 (30) | 79 (25) | 253 (32) | 68 (36) | 234 (29) | 30 (28) | 135 (27) | 197 (33) | 120 (29) | 212 (31) |
| *Late in the month (last week)* | 171 (16) | 61 (20) | 110 (14) | 25 (13) | 134 (17) | 12 (11) | 79 (16) | 92 (16) | 65 (16) | 106 (16) |
| *Middle of the month* | 169 (15) | 45 (14) | 124 (16) | 30 (16) | 122 (15) | 17 (16) | 67 (13) | 102 (17) | 67 (16) | 102 (15) |
| *No preference* | 426 (39) | 127 (41) | 299 (38) | 65 (35) | 311 (39) | 50 (46) | 225 (44) | 201 (34) | 167 (40) | 259 (38) |
| Preferred time(s) of day to come to clinic visits |  |  |  |  |  |  |  |  |  |  |
| *Before work (before 8 am)* | 392 (36) | 96 (31) | 296 (38) | 53 (28) | 287 (36) | 52 (48) | 165 (33) | 227 (38) | 161 (38) | 231 (34) |
| *Mornings (8 am to 12 pm)* | 593 (54) | 171 (55) | 422 (54) | 108 (57) | 425 (53) | 60 (55) | 254 (50) | 339 (57) | 224 (53) | 369 (54) |
| *Lunch time (12 am to 2 pm)* | 101 (9) | 27 (9) | 74 (9) | 16 (9) | 73 (9) | 12 (11) | 38 (8) | 63 (11) | 32 (8) | 69 (10) |
| *Afternoons (2 to 4 pm)* | 104 (9) | 29 (9) | 75 (10) | 26 (14) | 68 (8) | 10 (9) | 42 (8) | 62 (10) | 50 (12) | 54 (8) |
| *After work (4-7 pm)* | 42 (4) | 15 (5) | 27 (3) | 10 (5) | 28 (3) | 4 (4) | 16 (3) | 26 (4) | 22 (5) | 20 (3) |
| *Weekends* | 50 (5) | 23 (7) | 27 (3) | 6 (3) | 37 (5) | 7 (6) | 30 (6) | 20 (3) | 23 (5) | 27 (4) |
| *Other* | 16 (1) | 6 (2) | 10 (1) | 3 (2) | 11 (1) | 2 (2) | 14 (3) | 2 (0) | 8 (2) | 8 (1) |
| Preference for bringing companion (friend, family, support person) to | 114 (10) | 31 (10) | 83 (11) | 23 (12) | 79 (10) | 12 (11) | 50 (10) | 64 (11) | 46 (11) | 68 (10) |
|  |  | Male | Female | 18-24 | 25-49 | 50+ | Rural | Urban | *I/RI | ≤6 mos |
| clinic (yes) |  |  |  |  |  |  |  |  |  |  |
| How many months of HIV medications did you receive today? |  |  |  |  |  |  |  |  |  |  |
| None | 31 (3) | 11(4) | 20 (3) | 7 (4) | 21 (3) | 3 (3) | 19 (4) | 12 (2) | 6 (1) | 25 (4) |
| < One month | 2 (0) | 1 (0) | 1 (0) | 0 (0) | 2 (0) | 0 (0) | 0 (0) | 2 (0) | 2 (0) | - |
| One month | 896 (82) | 241 (77) | 655 (83) | 156 (83) | 648 (81) | 92 (84) | 400 (79) | 496 (84) | 388 (93) | 508 (75) |
| Two months | 114 (10) | 42 (13) | 72 (9) | 15 (8) | 93 (12) | 6 (5) | 59 (12) | 55 (9) | 10 (2) | 104 (15) |
| Three months | 49 (4) | 14 (4) | 35 (4) | 10 (5) | 34 (4) | 5 (5) | 26 (5) | 23 (4) | 13 (3) | 36 (5) |
| Other | 6 (1) | 3 (1) | 3 (0) | 0 (0) | 3 (0) | 3 (3) | 2 (0) | 4 (1) |  | 6 (1) |
| Preference for how many months of medications to be dispensed |  |  |  |  |  |  |  |  |  |  |
| 1 month at a time | 125 (11) | 44 (14) | 81 (10) | 23 (12) | 89 (11) | 13 (12) | 59 (12) | 66 (11) | 61 (15) | 64 (9) |
| 2 months at a time | 276 (25) | 83 (27) | 193 (25) | 46 (24) | 199 (25) | 31 (28) | 128 (25) | 148 (25) | 108 (26) | 168 (25) |
| 3 months at a time | 487 (44) | 128 (41) | 359 (46) | 75 (40) | 362 (45) | 50 (46) | 231 (46) | 256 (43) | 183 (44) | 304 (45) |
| 4 months at a time | 16 (1) | 5 (2) | 11 (1) | 4 (2) | 11 (1) | 1 (1) | 8 (2) | 8 (1) | 7 (2) | 9 (1) |
| 6 months at a time | 194 (18) | 52 (17) | 142 (18) | 40 (21) | 140 (17) | 14 (13) | 80 (16) | 114 (19) | 60 (14) | 134 (20) |
| Preference for medication packaging |  |  |  |  |  |  |  |  |  |  |
| One bottle for each month | 478 (44) | 144 (46) | 334 (42) | 78 (41) | 346 (43) | 54 (50) | 230 (45) | 248 (42) | 196 (47) | 282 (42) |
| One larger bottle with several months in it | 170 (15) | 44 (14) | 126 (16) | 28 (15) | 129 (16) | 13 (12) | 70 (14) | 100 (17) | 60 (14) | 110 (16) |
| Unmarked (blank) container | 78 (7) | 20 (6) | 58 (7) | 17 (9) | 60 (7) | 1 (1) | 38 (8) | 40 (7) | 29 (7) | 49 (7) |
| Container with instruction label | 78 (7) | 27 (9) | 51 (6) | 12 (6) | 59 (7) | 7 (6) | 42 (8) | 36 (6) | 34 (8) | 44 (6) |
| Blister pack | 147 (13) | 33 (11) | 114 (15) | 31 (16) | 104 (13) | 12 (11) | 61 (12) | 86 (15) | 50 (12) | 97 (14) |
| Any kind of packaging is fine | 143 (13) | 40 (13) | 103 (13) | 21 (11) | 101 (13) | 21 (19) | 62 (12) | 81 (14) | 48 (11) | 95 (14) |
| Something else(specify) | 4 (0) | 4 (1) | 0 (0) | 1 (1) | 2 (0) | 1 (1) | 3 (1) | 1 (0) | 2 (0) | 2 (0) |
| Preferred provider and service location |  |  |  |  |  |  |  |  |  |  |
| Preference for healthcare provider cadre |  |  |  |  |  |  |  |  |  |  |
| Doctor or clinical officer | 160 (15) | 64 (21) | 96 (12) | 17 (9) | 126 (16) | 17 (16) | 60 (12) | 100 (17) | 69 (16) | 91 (13) |
|  |  | Male | Female | 18-24 | 25-49 | 50+ | Rural | Urban | *I/RI | ≤6 mos |
| Nurse | 872 (79) | 237 (76) | 635 (81) | 160 (85) | 625 (78) | 87 (80) | 421 (83) | 451 (76) | 325 (78) | 547 (81) |
| Counsellor | 52 (5) | 7 (2) | 45 (6) | 10 (5) | 38 (5) | 4 (4) | 19 (4) | 33 (6) | 21 (5) | 31 (5) |
| CHW, peer/expert patient | 7 (1) | 3 (1) | 4 (1) | 0 (0) | 7 (1) | 0 (0) | 4 (1) | 3 (1) | 4 (1) | 10 (1) |
| Someone else (specify) | 7 (1) | 1 (0) | 6 (1) | 1 (1) | 5 (1) | 1 (1) | 2 (0) | 5 (1) | 1 (0) | 6 (1) |
| Preferred service location |  |  |  |  |  |  |  |  |  |  |
| Community-based (e.g. at a school or church or pharmacy) | 710 (65) | 206 (66) | 504 (64) | 113 (60) | 534 (67) | 63 (58) | 300 (59) | 410 (69) | 268 (64) | 442 (65) |
| Home medication delivery | 580 (53) | 168 (54) | 412 (52) | 94 (50) | 421 (53) | 65 (60) | 240 (47) | 340 (57) | 232 (55) | 348 (51) |
| **Counselling and education preferences** |  |  |  |  |  |  |  |  |  |  |
| Preference for quantity of one-on-one counselling to help manage your treatment, compared to what you received |  |  |  |  |  |  |  |  |  |  |
| More | 535 (49) | 153 (49) | 382 (49) | 89 (47) | 390 (49) | 56 (51) | 255 (50) | 280 (47) | 218 (52) | 317 (47) |
| Same | 540 (49) | 150 (48) | 390 (50) | 92 (49) | 396 (49) | 52 (48) | 242 (48) | 298 (50) | 191 (46) | 349 (51) |
| Less | 23 (2) | 9 (3) | 14 (2) | 7 (4) | 15 (2) | 1 (1) | 9 (2) | 14 (2) | 10 (2) | 13 (2) |
| Preference for quantity of information and education about HIV and ART, compared to what you received |  |  |  |  |  |  |  |  |  |  |
| More | 530 (48) | 150 (48) | 380 (48) | 89 (47) | 393 (49) | 48 (44) | 254 (50) | 276 (47) | 202 (48) | 328 (48) |
| Same | 538 (49) | 155 (50) | 383 (49) | 90 (48) | 390 (49) | 58 (53) | 246 (49) | 292 (49) | 205 (49) | 333 (49) |
| Less | 30 (3) | 7 (2) | 23 (3) | 9 (5) | 18 (2) | 3 (3) | 6 (1) | 24 (4) | 12 (3) | 18 (3) |
| Preference for format to receive information about HIV and ART** |  |  |  |  |  |  |  |  |  |  |
| Written material (brochure or information sheet) | 401 (37) | 129 (41) | 272 (35) | 59 (31) | 293 (37) | 49 (45) | 213 (42) | 188 (32) | 143 (34) | 258 (38) |
| Class/group session in community (not at clinic) | 89 (8) | 13 (4) | 76 (10) | 22 (12) | 57 (7) | 10 (9) | 23 (5) | 66 (11) | 45 (11) | 44 (6) |
|  |  | Male | Female | 18-24 | 25-49 | 50+ | Rural | Urban | *I/RI | ≤6 mos |
| *Class/group session with provider at clinic* | 191 (17) | 51 (16) | 140 (18) | 20 (11) | 140 (17) | 31 (28) | 85 (17) | 106 (18) | 95 (23) | 96 (14) |
| *One-on-one session with provider at clinic* | 543 (49) | 165 (53) | 378 (48) | 93 (49) | 393 (49) | 57 (52) | 262 (52) | 281 (47) | 217 (52) | 326 (48) |
| *Social media (e.g. Facebook, Twitter)* | 241 (22) | 60 (19) | 181 (23) | 59 (31) | 171 (21) | 11 (10) | 68 (13) | 173 (29) | 99 (24) | 142 (21) |
| *Group within my community* | 43 (4) | 10 (3) | 33 (4) | 5 (3) | 32 (4) | 6 (6) | 16 (3) | 27 (5) | 22 (5) | 21 (3) |
| *Radio or TV* | 262 (24) | 76 (24) | 186 (24) | 33 (18) | 187 (23) | 42 (39) | 103 (20) | 159 (27) | 111 (26) | 151 (22) |
| *Videos to watch online at home* | 112 (10) | 28 (9) | 84 (11) | 25 (13) | 81 (10) | 6 (6) | 37 (7) | 75 (13) | 42 (10) | 70 (10) |
| *Text messages on my phone* | 589 (54) | 165 (53) | 424 (54) | 102 (54) | 435 (54) | 52 (48) | 293 (58) | 296 (50) | 215 (51) | 374 (55) |
| *Links to websites that I can browse in my own time* | 194 (18) | 47 (15) | 147 (19) | 50 (27) | 137 (17) | 7 (6) | 89 (18) | 105 (18) | 68 (16) | 126 (19) |
| *Other* | 3 (0) | 1 (0) | 2 (0) | 0 (0) | 3 (0) | 0 (0) | 1 (0) | 2 (0) | 2 (0) | 1 (0) |
| Preferred language for receiving information about HIV and ART |  |  |  |  |  |  |  |  |  |  |
| *English* | 422 (38) | 113 (36) | 309 (39) | 104 (55) | 297 (37) | 21 (19) | 147 (29) | 275 (46) | 166 (40) | 256 (38) |
| *isiZulu* | 301 (27) | 92 (29) | 209 (27) | 42 (22) | 218 (27) | 41 (38) | 202 (40) | 99 (17) | 79 (19) | 222 (33) |
| *Xitsonga* | 74 (7) | 26 (8) | 48 (6) | 9 (5) | 60 (7) | 5 (5) | 28 (6) | 46 (8) | 40 (10) | 34 (5) |
| *Siswati* | 165 (15) | 47 (15) | 118 (15) | 17 (9) | 129 (16) | 19 (17) | 87 (17) | 78 (13) | 82 (20) | 83 (12) |

\*I/RI = Initiating ART on the day of PREFER study enrollment; ≤ 6 mos = On ART for 6 months or less at time of study enrollment
\*\*Clients could select as many as apply

**Supplementary Table 3.** Participants’ preferences for treatment in the first six months by sex, age, and time on ART at enrolment, Zambia.

| Preference (n, %) | Total | Sex |  | Age groups in years |  |  | Place of residence |  | Time on ART at study enrolment |  |
| --- | --- | --- | --- | --- | --- | --- | --- | --- | --- | --- |
|  |  | Male | Female | 18-24 | 25-49 | 50+ | Rural | Urban | I/RI | ≤6 months |
| N | 771 | 257 | 514 | 123 | 596 | 52 | 623 | 148 | 261 | 510 |
| ***Preferred visit and dispensing procedures*** |  |  |  |  |  |  |  |  |  |  |
| Offered choice about service delivery since starting ART (yes) | 100 (13) | 30 (12) | 70 (14) | 16 (13) | 79 (13) | 5 (10) | 83 (13) | 17 (11) | 31 (12) | 69 (14) |
| Preference for clinic visit frequency |  |  |  |  |  |  |  |  |  |  |
| *Every month* | 61 (8) | 22 (9) | 39 (8) | 11 (9) | 47 (8) | 3 (6) | 47 (8) | 14 (9) | 34 (13) | 27 (5) |
| *Every 2 months* | 72 (9) | 33 (13) | 39 (8) | 14 (11) | 55 (9) | 3 (6) | 48 (8) | 24 (16) | 32 (12) | 40 (8) |
| *Every 3 months* | 233 (30) | 75 (29) | 158 (31) | 34 (28) | 177 (30) | 22 (42) | 187 (30) | 46 (31) | 77 (30) | 156 (31) |
| *Every 6 months* | 372 (48) | 115 (45) | 257 (50) | 56 (46) | 292 (49) | 24 (46) | 313 (50) | 59 (40) | 103 (39) | 269 (53) |
| *Other* | 33 (4) | 12 (5) | 21 (4) | 8 (7) | 25 (4) | 0 (0) | 28 (4) | 5 (3) | 15 (6) | 18 (4) |
| Preferred part of the month to visit clinic |  |  |  |  |  |  |  |  |  |  |
| *Early in the month (first week)* | 313 (41) | 99 (39) | 214 (42) | 55 (45) | 238 (40) | 20 (38) | 242 (39) | 71 (48) | 105 (40) | 208 (41) |
| *Late in the month (last week)* | 187 (24) | 65 (25) | 122 (24) | 28 (23) | 143 (24) | 16 (31) | 158 (25) | 29 (20) | 62 (24) | 125 (25) |
| *Middle of the month* | 96 (12) | 30 (12) | 66 (13) | 18 (15) | 77 (13) | 1 (2) | 84 (13) | 12 (8) | 35 (13) | 61 (12) |
| *No preference* | 175 (23) | 63 (25) | 112 (22) | 22 (18) | 138 (23) | 15 (29) | 139 (22) | 36 (24) | 59 (23) | 116 (23) |
| Preferred time(s) of day to come to clinic visits |  |  |  |  |  |  |  |  |  |  |
| *Before work (before 8 am)* | 202 (26) | 72 (28) | 130 (25) | 27 (22) | 158 (27) | 17 (33) | 146 (23) | 56 (38) | 70 (27) | 132 (26) |
| *Mornings (8 am to 12 pm)* | 429 (56) | 132 (51) | 297 (58) | 67 (54) | 328 (55) | 34 (65) | 346 (56) | 83 (56) | 133 (51) | 296 (58) |
| *Lunch time (12 am to 2 pm)* | 41 (5) | 10 (4) | 31 (6) | 6 (5) | 35 (6) | 0 (0) | 32 (5) | 9 (6) | 12 (5) | 29 (6) |
| *Afternoons (2 to 4 pm)* | 75 (10) | 33 (13) | 42 (8) | 9 (7) | 63 (11) | 3 (6) | 64 (10) | 11 (7) | 29 (11) | 46 (9) |
| *After work (4-7 pm)* | 58 (8) | 21 (8) | 37 (7) | 17 (14) | 40 (7) | 1 (2) | 53 (9) | 5 (3) | 26 (10) | 32 (6) |
| *Weekends* | 15 (2) | 5 (2) | 10 (2) | 4 (3) | 11 (2) | 0 (0) | 15 (2) | 0 (0) | 7 (3) | 8 (2) |
| *Other* | 19 (2) | 5 (2) | 14 (3) | 5 (4) | 14 (2) | 0 (0) | 19 (3) | 0 (0) | 6 (2) | 13 (3) |
| Preference for bringing companion (friend, family, support person) to clinic (yes) | 151 (20) | 57 (22) | 94 (18) | 28 (23) | 114 (19) | 9 (17) | 114 (18) | 37 (25) | 63 (24) | 88 (17) |
| How many months of HIV medications did you receive today? |  |  |  |  |  |  |  |  |  |  |
|  |  | Male | Female | 18-24 | 25-49 | 50+ | Rural | Urban | I/RI | ≤6 months |
| *None* | 34 (4) | 11 (4) | 23 (4) | 8 (7) | 24 (4) | 2 (4) | 24 (4) | 10 (7) | 8 (3) | 26 (5) |
| *< One month* | 28 (4) | 12 (5) | 16 (3) | 4 (3) | 22 (4) | 2 (4) | 23 (4) | 5 (3) | 23 (9) | 5 (1) |
| *One month* | 274 (36) | 93 (36) | 181 (35) | 52 (42) | 206 (35) | 16 (31) | 219 (35) | 55 (37) | 170 (65) | 104 (20) |
| *Two months* | 59 (8) | 23 (9) | 36 (7) | 11 (9) | 43 (7) | 5 (10) | 38 (6) | 21 (14) | 6 (2) | 53 (10) |
| *Three months* | 334 (43) | 95 (37) | 239 (47) | 42 (34) | 267 (45) | 25 (48) | 284 (46) | 50 (34) | 45 (17) | 289 (57) |
| *Four months* | 4 (0) | 3 (1) | 1 (0) | 1 (1) | 3 (1) | 0 (0) | 2 (0) | 2 (1) | - | 4 (1) |
| *Six months* | 12 (2) | 6 (2) | 6 (1) | 0 (0) | 11 (2) | 1 (2) | 10 (2) | 2 (1) | 2 (1) | 10 (2) |
| *Other* | 26 (3) | 14 (5) | 12 (2) | 5 (4) | 20 (3) | 1 (2) | 23 (4) | 3 (2) | 7 (3) | 19 (4) |
| Preference for how many months of medications to be dispensed |  |  |  |  |  |  |  |  |  |  |
| *1 month at a time* | 39 (5) | 19 (7) | 20 (4) | 12 (10) | 26 (4) | 1 (2) | 32 (5) | 7 (5) | 26 (10) | 13 (3) |
| *2 months at a time* | 59 (8) | 29 (11) | 30 (6) | 9 (7) | 48 (8) | 2 (4) | 44 (7) | 15 (10) | 26 (10) | 33 (6) |
| *3 months at a time* | 237 (31) | 78 (30) | 159 (31) | 30 (24) | 185 (31) | 22 (42) | 194 (31) | 43 (29) | 81 (31) | 156 (31) |
| *4 months at a time* | 17 (2) | 7 (3) | 10 (2) | 5 (4) | 11 (2) | 1 (2) | 12 (2) | 5 (3) | 6 (2) | 11 (2) |
| *6 months at a time* | 419 (54) | 124 (48) | 295 (57) | 67 (54) | 326 (55) | 26 (50) | 341 (55) | 78 (53) | 122 (47) | 297 (58) |
| Preference for medication packaging |  |  |  |  |  |  |  |  |  |  |
| *One bottle for each month* | 270 (35) | 93 (36) | 177 (34) | 32 (26) | 219 (37) | 19 (37) | 206 (33) | 64 (43) | 87 (33) | 183 (36) |
| *One larger bottle with several months in it* | 277 (36) | 85 (33) | 192 (37) | 44 (36) | 210 (35) | 23 (44) | 227 (36) | 50 (34) | 79 (30) | 198 (39) |
| *Unmarked (blank) container* | 53 (7) | 12 (5) | 41 (8) | 21 (17) | 30 (5) | 2 (4) | 45 (7) | 8 (5) | 21 (8) | 32 (6) |
| *Container with instruction label* | 84 (11) | 31 (12) | 53 (10) | 12 (10) | 71 (12) | 1 (2) | 68 (11) | 16 (11) | 38 (15) | 46 (9) |
| *Blister pack* | 41 (5) | 17 (7) | 24 (5) | 8 (7) | 30 (5) | 3 (6) | 38 (6) | 3 (2) | 23 (9) | 18 (4) |
| *Any kind of packaging is fine* | 41 (5) | 17 (7) | 24 (5) | 3 (2) | 34 (6) | 4 (8) | 34 (5) | 7 (5) | 12 (5) | 29 (6) |
| *Something else(specify)* | 5 (1) | 2 (1) | 3 (1) | 3 (2) | 2 (0) | 0 (0) | 5 (1) | 0 (0) | 1 (0) | 4 (1) |
| **Preferred provider and service location** |  |  |  |  |  |  |  |  |  |  |
| Preference for healthcare provider |  |  |  |  |  |  |  |  |  |  |
| *Doctor or clinical officer* | 382 (50) | 148 (58) | 234 (46) | 63 (51) | 292 (49) | 27 (52) | 308 (49) | 74 (50) | 122 (47) | 260 (51) |
| *Nurse* | 200 (26) | 55 (21) | 145 (28) | 30 (24) | 154 (26) | 16 (31) | 166 (27) | 34 (23) | 62 (24) | 138 (27) |
| *Counsellor* | 158 (20) | 43 (17) | 115 (22) | 25 (20) | 126 (21) | 7 (13) | 120 (19) | 38 (26) | 71 (27) | 87 (17) |
| *CHW, peer/expert patient* | 14 (2) | 4 (2) | 10 (2) | 4 (3) | 9 (2) | 1 (2) | 13 (2) | 1 (1) | 2 (1) | 12 (2) |
|  |  | Male | Female | 18-24 | 25-49 | 50+ | Rural | Urban | I/RI | ≤6 months |
| *Someone else (specify)* | 17 (2) | 7 (3) | 10 (2) | 1 (1) | 15 (3) | 1 (2) | 16 (3) | 1 (1) | 4 (2) | 13 (3) |
| Preference for service delivery location |  |  |  |  |  |  |  |  |  |  |
| *Community-based (e.g. at a school or church or pharmacy)* | 232 (30) | 77 (30) | 155 (30) | 30 (24) | 196 (33) | 6 (12) | 196 (31) | 36 (24) | 76 (29) | 156 (31) |
| *Home medication delivery* | 439 (57) | 151 (59) | 288 (56) | 64 (52) | 350 (59) | 25 (48) | 344 (55) | 95 (64) | 155 (59) | 284 (56) |
| **Counselling and education preferences** |  |  |  |  |  |  |  |  |  |  |
| Preference for quantity of one-on-one counselling to help manage your treatment, compared to what you received |  |  |  |  |  |  |  |  |  |  |
| *More* | 372 (48) | 118 (46) | 254 (49) | 68 (55) | 279 (47) | 25 (48) | 290 (47) | 82 (55) | 129 (49) | 243 (48) |
| *Same* | 369 (48) | 131 (51) | 238 (46) | 50 (41) | 294 (49) | 25 (48) | 307 (49) | 62 (42) | 124 (48) | 245 (48) |
| *Less* | 30 (4) | 8 (3) | 22 (4) | 5 (4) | 23 (4) | 2 (4) | 26 (4) | 4 (3) | 8 (3) | 22 (4) |
| Preference for quantity of information and education about HIV and ART, compared to what you received |  |  |  |  |  |  |  |  |  |  |
| *More* | 372 (48) | 114 (44) | 258 (50) | 72 (59) | 276 (46) | 24 (46) | 300 (48) | 72 (49) | 130 (50) | 242 (47) |
| *The same* | 369 (48) | 136 (53) | 233 (45) | 45 (37) | 298 (50) | 26 (50) | 300 (48) | 69 (47) | 122 (47) | 247 (48) |
| *Less* | 30 (4) | 7 (3) | 23 (4) | 6 (5) | 22 (4) | 2 (4) | 23 (4) | 7 (5) | 9 (3) | 21 (4) |
| Preference for format to receive information about HIV and ART** |  |  |  |  |  |  |  |  |  |  |
| *Written material (brochure or information sheet)* | 171 (22) | 61 (24) | 110 (21) | 33 (27) | 123 (21) | 15 (29) | 137 (22) | 34 (23) | 67 (26) | 104 (20) |
| *Class/group session in community (not at clinic)* | 51 (7) | 15 (6) | 36 (7) | 10 (8) | 36 (6) | 5 (10) | 38 (6) | 13 (9) | 20 (8) | 31 (6) |
| *Class/group session with provider at clinic* | 131 (17) | 34 (13) | 97 (19) | 29 (24) | 95 (16) | 7 (13) | 112 (18) | 19 (13) | 35 (13) | 96 (19) |
| *One-on-one session with provider at clinic* | 350 (45) | 116 (45) | 234 (46) | 56 (46) | 271 (45) | 23 (44) | 258 (41) | 92 (62) | 114 (44) | 236 (46) |
| *Social media (e.g. Facebook, Twitter)* | 55 (7) | 24 (9) | 31 (6) | 9 (7) | 44 (7) | 2 (4) | 45 (7) | 10 (7) | 21 (8) | 34 (7) |
| *Group within my community* | 19 (2) | 5 (2) | 14 (3) | 5 (4) | 13 (2) | 1 (2) | 14 (2) | 5 (3) | 8 (3) | 11 (2) |
|  |  | Male | Female | 18-24 | 25-49 | 50+ | Rural | Urban | I/RI | ≤6 months |
| *Radio or TV* | 183 (24) | 63 (25) | 120 (23) | 26 (21) | 139 (23) | 18 (35) | 146 (23) | 37 (25) | 63 (24) | 120 (24) |
| *Videos to watch online at home* | 13 (2) | 4 (2) | 9 (2) | 2 (2) | 11 (2) | 0 (0) | 13 (2) | 0 (0) | 2 (1) | 11 (2) |
| *Text messages on my phone* | 236 (31) | 93 (36) | 143 (28) | 34 (28) | 191 (32) | 11 (21) | 185 (30) | 51 (34) | 84 (32) | 152 (30) |
| *Links to websites that I can browse in my own time* | 6 (1) | 2 (1) | 4 (1) | 2 (2) | 3 (1) | 1 (2) | 5 (1) | 1 (1) | 2 (1) | 4 (1) |
| *Other* | 4 (1) | 1 (0) | 3 (1) | 0 (0) | 4 (1) | 0 (0) | 4 (1) | 0 (0) | 3 (1) | 1 (0) |
| Preferred language for receiving information about HIV and ART |  |  |  |  |  |  |  |  |  |  |
| *English* | 128 (17) | 48 (19) | 80 (16) | 29 (24) | 89 (15) | 10 (19) | 114 (18) | 14 (9) | 41 (16) | 87 (17) |
| *Nyanja* | 346 (45) | 109 (42) | 237 (46) | 56 (46) | 274 (46) | 16 (31) | 262 (42) | 84 (57) | 107 (41) | 239 (47) |
| *Bemba* | 217 (28) | 71 (28) | 146 (28) | 27 (22) | 172 (29) | 18 (35) | 205 (33) | 12 (8) | 84 (32) | 133 (26) |
| *Tonga* | 55 (7) | 20 (8) | 35 (7) | 8 (7) | 42 (7) | 5 (10) | 26 (4) | 29 (20) | 20 (8) | 35 (7) |

\*I/RI = Initiating ART on the day of PREFER study enrollment; ≤ 6 mos = On ART for 6 months or less at time of study enrollment
\*\*Clients could select as many as apply

